# Adjuvant corticosteroid therapy for critically ill patients with COVID-19

**DOI:** 10.1101/2020.04.07.20056390

**Authors:** Xiaofan Lu, Taige Chen, Yang Wang, Jun Wang, Bing Zhang, Yongsheng Li, Fangrong Yan

## Abstract

Addition of adjuvant corticosteroid therapy to standard antiviral treatment of patients with coronavirus disease (COVID-19) is common in clinical practice. However, evidence is scarce regarding the efficacy of adjuvant corticosteroids in patients who are critically ill. We retrospectively evaluated the effects of adjuvant corticosteroid treatment on the outcome of 244 critically ill patients with COVID-19, using a risk stratification model that adjusts for potential differences between the steroid group (n=151) and the non-steroid group (n=93). We observed that adjuvant corticosteroid therapy was independent from 28-day mortality, either in multivariate logistic regression of the entire cohort after adjustment for major mortality-associated variables (age, SpO_2_/FiO_2_, and lymphocyte count) and individual propensity score (adjusted OR: 1.05; 95% CI: −1.92-2.01), or in propensity score-matched (1:1 without replacement) case-control analysis (62 patients in 31 pairs; log-rank test *P*=0.17). Additionally, subgroup analyses of 147 (60%) patients who had dyspnea and 87 (36%) patients who had ARDS revealed corticosteroid treatment was not associated with clinical outcome (both, *P*>0.3). However, increased corticosteroids dosage was significantly associated with elevated mortality risk after adjustment for administration duration (*P*=0.003); every ten-milligram increase in hydrocortisone-equivalent dosage was associated with additional 4% mortality risk (adjusted HR: 1.04, 95% CI: 1.01-1.07). Our findings indicated that limited effect of corticosteroid therapy could pose to overall survival and prudent dose within effective limits may be recommended for critically ill patients under certain circumstances.

## Introduction

The spread of coronavirus disease (COVID-19) has already taken on pandemic proportions. A substantial portion of patients developed rapidly progressive pneumonia leading to acute respiratory distress syndrome (ARDS) and multiple organ dysfunctions, conditions associated with high mortality [1, 2]. Adjuvant corticosteroid therapy of such patients is common in clinical practice. However, evidence is scarce regarding the efficacy of adjuvant corticosteroids in patients who are critically ill with COVID-19. We retrospectively evaluated the effects of adjuvant corticosteroid treatment on the outcome of critically ill patients with COVID-19, using a risk stratification model that adjusts for potential differences between the steroid group and the non-steroid group.

## Methods

### Study population

We retrospectively reviewed medical records of all adult patients with confirmed infection of severe acute respiratory syndrome coronavirus 2 (SARS-CoV2) who were admitted to intensive care wards of Tongji hospital in Wuhan, China, from January 25 through February 25, 2020. A total of 244 eligible patients who had complete medical records and were at least 20 years old, critically ill, and treated with etiological antiviral agents were enrolled. Critically ill patients were defined as those who were admitted to intensive care wards and required mechanical ventilation (either invasive or noninvasive), or with ARDS (PaO_2_/FiO_2_ ≤300mmHg; when PaO_2_ is not available, SpO_2_/FiO_2_ ≤315 suggests ARDS), or sepsis with acute organ dysfunction [3]. The clinical outcome was 28-day mortality after admission. The Ethics Commission of Tongji hospital approved this study, with a waiver of informed consent.

### Study design

The first part of this study was a retrospective cohort study that included all eligible patients. The outcome of patients given corticosteroids was compared with those not given. The second part was a 1:1 matched case-control study. The patients with corticosteroids treatment (*e.g*., methylprednisolone, dexamethasone, and hydrocortisone) were designated as “case subjects”, and those without as “control subjects”. We converted all preparations to hydrocortisone-equivalent doses (methylprednisolone 1:5, dexamethasone 1:25) [4].

### Statistical analyses

To reduce the effect of corticosteroids treatment selection bias and potential confounding, we adjusted for differences in baseline characteristics by propensity score, which was estimated to predict a patient’s probability of being treated with a corticosteroid without regard to outcomes, using multivariate logistic regression [5]. Potential confounders considered in propensity score matching (PSM) analysis were those variables included in the final model by means of step-wise backward elimination procedures with *P*<0.20 [6]. Model discrimination and calibration were assessed with c-statistics (*c*=0.94), and Hosmer-Lemeshow goodness of fit test (*P*=0.93), respectively. The effect of corticosteroids treatment on clinical outcome was analyzed by multivariate logistic regression with adjustment for major variables (age, SpO_2_/FiO_2_, and lymphocyte count) associated with mortality; the individual propensity score was incorporated into the model as a covariable to calculate the propensity adjusted odds ratio (OR) [6]. Additionally, PSM was performed to generate propensity score-matched pairs without replacement, and the survival probability was compared between matched pairs by Kaplan-Meier curve and analyzed with log-rank test. Cox regression was used to estimate hazard ratio (HR) with 95% CI. Continuous variables were described as the median (IQR) while categorical variables were expressed as frequencies (%). All statistical analyses were conducted with R (version 3.6.2) using a Fisher’s exact test for categorical data and Mann-Whitney test for continuous data. For unadjusted comparisons, a two-sided *P*<0.05 was considered statistically significant.

## Results

### Cohort study

Of the 244 critically ill patients with confirmed SARS-CoV2, the median age was 62 (50-71) years, and 52% were male (Table). All patients were given antiviral therapy (e.g., oseltamivir, arbidol, lopinavir/ritonavir, ganciclovir, interferon-α), and 151 (62%) were given adjuvant corticosteroid treatment (hydrocortisone-equivalent dosage range: 100-800mg/d). The median (IQR) administration duration of corticosteroid treatment was 8 (4-12) days. Multiple organ dysfunctions were more common in the steroid group than in the non-steroid group. Multivariate analysis that adjusted for major mortality-associated variables and propensity score indicated that corticosteroid treatment was independent from overall mortality (adjusted OR: 1.05; 95% CI: −1.92-2.01). Of the 244 patients in this study, 147 (60%) had dyspnea and 87 (36%) had ARDS, and in subgroup analyses, multivariate analysis after adjustment revealed corticosteroid treatment was not significantly associated with increased 28-day mortality (both, *P*>0.3).

### Matched case-control study

Sixty-two patients in 31 pairs with and without corticosteroid therapy were matched by PSM with balanced baseline characteristics (all, P>0.05; Table). The 28-day mortality rate was 39% (12 out of 31) in case subjects and 16% (5 out of 31) in control subjects (*P*=0.09). Likewise, we did not observe statistical significance in survival probability of this matched case-control study, stratified by corticosteroid treatment (*P*=0.17; Figure). Unexpectedly, increased corticosteroids dosage was significantly associated with elevated mortality risk (*P*=0.003) in matched cases after adjustment for administration duration; every ten-milligram increase in hydrocortisone dosage was associated with additional 4% mortality risk (adjusted HR: 1.04, 95% CI: 1.01-1.07).

## Discussion

Our investigation indicated that addition of adjuvant corticosteroid therapy to standard antiviral treatment of critically ill patients with COVID-19 was not associated with the 28-day mortality, which means limited effect of corticosteroid therapy could pose to overall survival of such disease.

Given the various adverse effects (*e.g*., avascular necrosis, diabetes, delayed virus clearance, increased risk of secondary infections), corticosteroid therapy must be commenced with caution after full consideration of indications and contraindications, and routine corticosteroid use should be avoided during the process of anti-infection. Additionally, we observed that increased corticosteroids dosage was associated with elevated mortality risk, suggesting prudent dosage of corticosteroid should be promoted when necessary. We acknowledged limitations of the study. First, we did not distinguish patients who received corticosteroids for their underlying disease (*e.g*., COPD), the number of which was however relatively small. Second, PSM is limited by adjusting for observed covariables only, but in randomized trials randomization balances for known and unknown factors. Owing to this limitation, randomized placebo-controlled trials are warranted to resolve this clinically essential issue.

Altogether, abuse of corticosteroids is highly opposed and this study provided evidence that low-dose corticosteroids within effective limits may be recommended for critically ill patients with COVID-19 under certain circumstances.

## Data Availability

Some or all data generated or used during the study are available from the corresponding author by request. Dr J. Wang had full access to all of the data in the study. After publication, the data will be made available to others on reasonable requests after approval from the author (J.W) and Wuhan Tongji Hospital.

https://pan.baidu.com/s/1ujFeZUk7hBBIoEnF2uD-xg

## Acknowledgments

We would like to thank all the hospital staff members for their efforts in collecting the information that was used in this study, and all the patients who consented to donate their data for analysis and the medical staff members who are on the frontline of caring for patients. This work was supported by the National Key R&D Program of China (2019YFC1711000), the National Natural Science Foundation of China (81973145), the “Double First-Class” University Project (CPU2018GY09), the China Postdoctoral Science Foundation (2019M651805), the Science Foundation of Jiangsu Commission of Health (H2018117), and the Emergency Project for the Prevention and Control of the Novel Coronavirus Outbreak in Suzhou (SYS2020012).

## Author Contributions

Conceptualization: X. Lu, T. Chen.

Acquisition, analysis, or interpretation of data: J. Wang, Y. Wang, X. Lu, T. Chen.

Statistical analysis: X. Lu, F. Yan.

Investigation: X. Lu, T. Chen, Y. Wang, Y. Li, B. Zhang.

Drafting of the manuscript editing: X. Lu, T. Chen, Y. Wang.

Funding acquisition: J. Wang, Y. Wang, F. Yan.

Supervision: F. Yan.

## Data availability

Dr J. Wang had full access to all of the data in the study. After publication, the data will be made available to others on reasonable requests after approval from the author (J.W) and Wuhan Tongji Hospital.

---

**Table.**
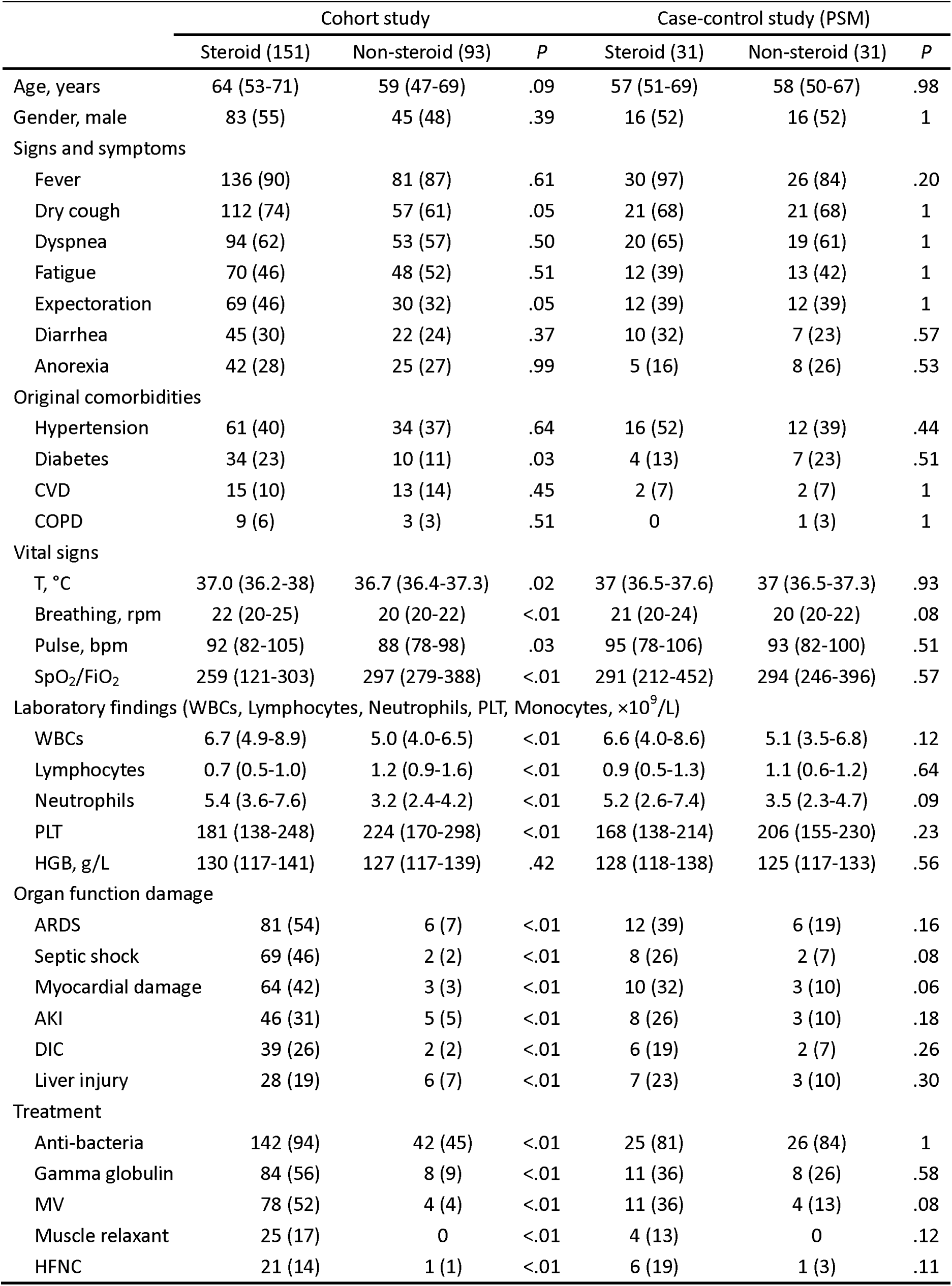
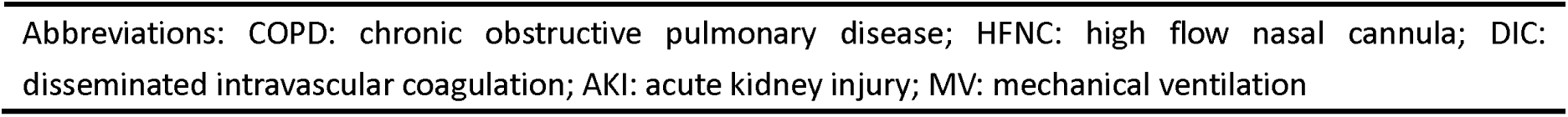
Baseline characteristics for steroid treatment and non-steroid treatment groups comprising critically ill patients with COVID-19 before and after propensity score matching

|  | Cohort study |  |  | Case-control study (PSM) |  |  |
| --- | --- | --- | --- | --- | --- | --- |
|  | Steroid (151) | Non-steroid (93) | *P* | Steroid (31) | Non-steroid (31) | *P* |
| Age, years | 64 (53-71) | 59 (47-69) | .09 | 57 (51-69) | 58 (50-67) | .98 |
| Gender, male | 83 (55) | 45 (48) | .39 | 16 (52) | 16 (52) | 1 |
| Signs and symptoms |  |  |  |  |  |  |
| Fever | 136 (90) | 81 (87) | .61 | 30 (97) | 26 (84) | .20 |
| Dry cough | 112 (74) | 57 (61) | .05 | 21 (68) | 21 (68) | 1 |
| Dyspnea | 94 (62) | 53 (57) | .50 | 20 (65) | 19 (61) | 1 |
| Fatigue | 70 (46) | 48 (52) | .51 | 12 (39) | 13 (42) | 1 |
| Expectoration | 69 (46) | 30 (32) | .05 | 12 (39) | 12 (39) | 1 |
| Diarrhea | 45 (30) | 22 (24) | .37 | 10 (32) | 7 (23) | .57 |
| Anorexia | 42 (28) | 25 (27) | .99 | 5 (16) | 8 (26) | .53 |
| Original comorbidities |  |  |  |  |  |  |
| Hypertension | 61 (40) | 34 (37) | .64 | 16 (52) | 12 (39) | .44 |
| Diabetes | 34 (23) | 10 (11) | .03 | 4 (13) | 7 (23) | .51 |
| CVD | 15 (10) | 13 (14) | .45 | 2 (7) | 2 (7) | 1 |
| COPD | 9 (6) | 3 (3) | .51 | 0 | 1 (3) | 1 |
| Vital signs |  |  |  |  |  |  |
| T, °C | 37.0 (36.2-38) | 36.7 (36.4-37.3) | .02 | 37 (36.5-37.6) | 37 (36.5-37.3) | .93 |
| Breathing, rpm | 22 (20-25) | 20 (20-22) | <.01 | 21 (20-24) | 20 (20-22) | .08 |
| Pulse, bpm | 92 (82-105) | 88 (78-98) | .03 | 95 (78-106) | 93 (82-100) | .51 |
| SpO<sub>2</sub>/FiO<sub>2</sub> | 259 (121-303) | 297 (279-388) | <.01 | 291 (212-452) | 294 (246-396) | .57 |
| Laboratory findings (WBCs, Lymphocytes, Neutrophils, PLT, Monocytes, ×10<sup>9</sup>/L) |  |  |  |  |  |  |
| WBCs | 6.7 (4.9-8.9) | 5.0 (4.0-6.5) | <.01 | 6.6 (4.0-8.6) | 5.1 (3.5-6.8) | .12 |
| Lymphocytes | 0.7 (0.5-1.0) | 1.2 (0.9-1.6) | <.01 | 0.9 (0.5-1.3) | 1.1 (0.6-1.2) | .64 |
| Neutrophils | 5.4 (3.6-7.6) | 3.2 (2.4-4.2) | <.01 | 5.2 (2.6-7.4) | 3.5 (2.3-4.7) | .09 |
| PLT | 181 (138-248) | 224 (170-298) | <.01 | 168 (138-214) | 206 (155-230) | .23 |
| HGB, g/L | 130 (117-141) | 127 (117-139) | .42 | 128 (118-138) | 125 (117-133) | .56 |
| Organ function damage |  |  |  |  |  |  |
| ARDS | 81 (54) | 6 (7) | <.01 | 12 (39) | 6 (19) | .16 |
| Septic shock | 69 (46) | 2 (2) | <.01 | 8 (26) | 2 (7) | .08 |
| Myocardial damage | 64 (42) | 3 (3) | <.01 | 10 (32) | 3 (10) | .06 |
| AKI | 46 (31) | 5 (5) | <.01 | 8 (26) | 3 (10) | .18 |
| DIC | 39 (26) | 2 (2) | <.01 | 6 (19) | 2 (7) | .26 |
| Liver injury | 28 (19) | 6 (7) | <.01 | 7 (23) | 3 (10) | .30 |
| Treatment |  |  |  |  |  |  |
| Anti-bacteria | 142 (94) | 42 (45) | <.01 | 25 (81) | 26 (84) | 1 |
| Gamma globulin | 84 (56) | 8 (9) | <.01 | 11 (36) | 8 (26) | .58 |
| MV | 78 (52) | 4 (4) | <.01 | 11 (36) | 4 (13) | .08 |
| Muscle relaxant | 25 (17) | 0 | <.01 | 4 (13) | 0 | .12 |
| HFNC | 21 (14) | 1 (1) | <.01 | 6 (19) | 1 (3) | .11 |

Abbreviations: COPD: chronic obstructive pulmonary disease; HFNC: high flow nasal cannula; DIC: disseminated intravascular coagulation; AKI: acute kidney injury; MV: mechanical ventilation

**Figure.**
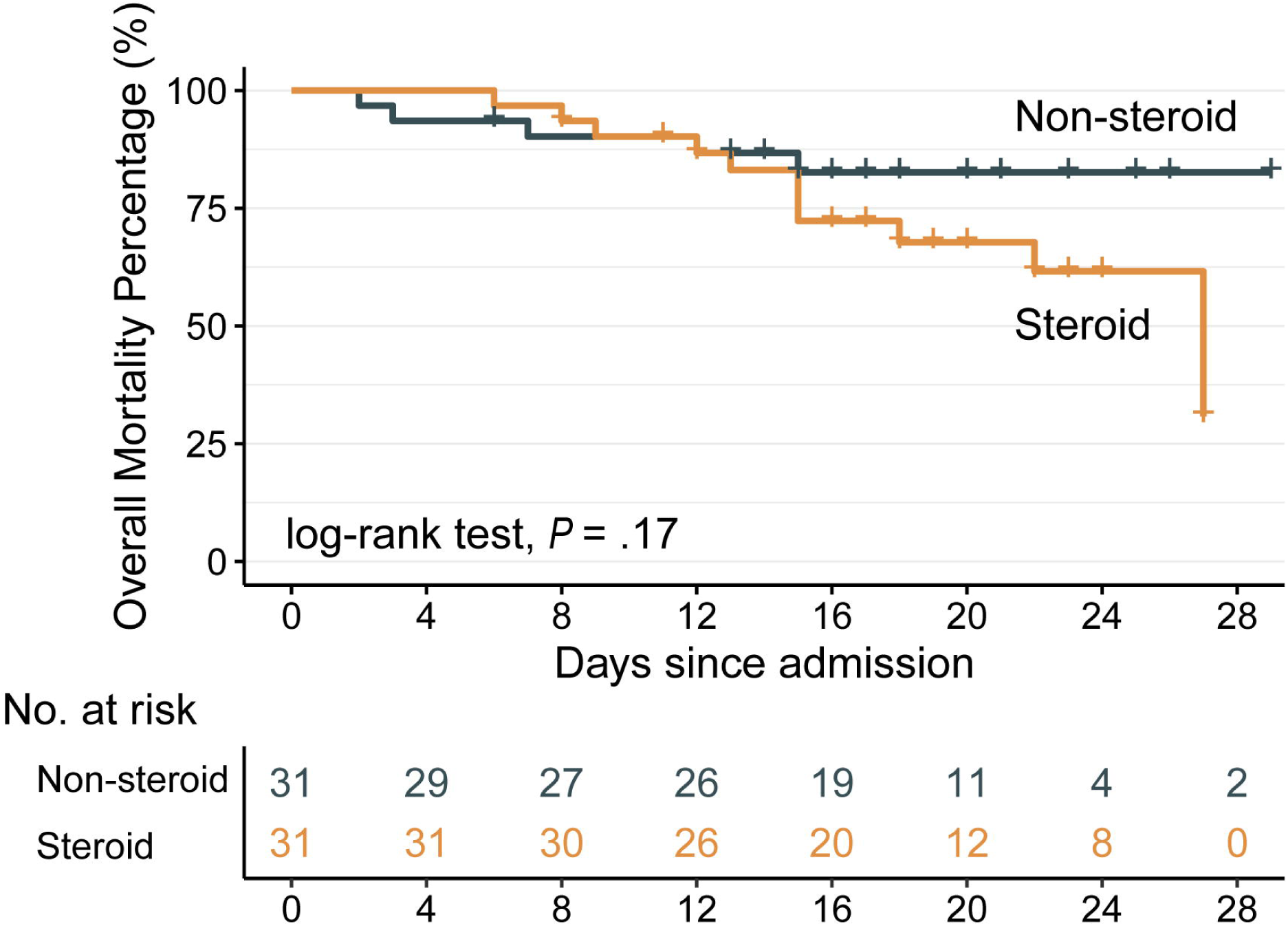
Survival curves stratified by adjuvant corticosteroid treatment. Thirty-one critically ill patients with COVID-19 who received corticosteroid treatment (yellow line) are compared with 31 matched control subjects (green line) who did not receive corticosteroid treatment.

